# Combination AI-Machine Learning to Diagnose Pulmonary Hypertension: A Real-World Evidence Cohort Study

**DOI:** 10.1101/2025.09.30.25336749

**Authors:** Seyed M. Shams, Mary E. Maldarelli, Steven Cassady, Gautam Ramani, Colleen M. Ennett, Bradley A. Maron, Katarina Zeder

## Abstract

**BACKGROUND:** Pulmonary hypertension (PH) is a highly morbid disease, but underdiagnosis is common outside of expert referral centers. Consequentially, there may be opportunities to automate PH diagnosis using artificial intelligence (AI) clinical decision support tools. Analysis of patient-level right heart catheterization (RHC) data is required to optimize AI-based PH diagnosis but has not been reported previously.

**METHODS:** We performed a retrospective cohort analysis of all RHC studies (January 1, 2016 to December 31, 2024) performed at the University of Maryland Medical System (UMMS), which is a Maryland statewide clinical network of 12 hospitals serving >2 million patients. We developed an automated large language model (LLM)-driven Pattern Repository (LDPR) method, featuring three task-specific LLM agents for extracting unstructured RHC data, which was manually cross-validated independently by two PH experts. To address data missingness, we used machine-learning to develop formulae to calculate mean pulmonary artery pressure (mPAP) from systolic (sPAP) and diastolic (dPAP) PAP, using an 80/20 train-test split.

**RESULTS:** The study cohort included N=11,029 unique patients and 17,292 RHC reports (age 66±13.5 years; 43% female; 65% White, 30% Black or African American; mPAP, 28±11mmHg; 26% congestive heart failure). The precision for accurate mPAP, sPAP, and dPAP extraction by the LLM was 99.6%, 99.4%, and 99.4%, respectively, with a detection failure of 0.4%. A missing mPAP was noted in N=548 cases and N=507 unique patients (3.2% and 4.6%, respectively). When applying ML to the dataset, the simple, linear equation: mPAP=1.51+0.43*sPAP+0.45*dPAP returned the highest R2 of 0.94 and lowest mean square error of 8.3 mmHg, which outperformed linear equations used currently (all p<0.001). The ML-derived formula was then directed to patients with missing mPAP (N=507) and identified N=382 patients (75.3%) with mPAP >20mmHg, and therefore reclassifying patients from no diagnosis to a diagnosis of PH.

**CONCLUSION:** In this retrospective cohort analysis, combination LLM-ML-based extraction and interpretation of RHC was used to automate PH diagnosis in a large and heterogenous patient population. This approach is an efficient and scalable solution to preventing under-diagnosis of PH and demonstrates the feasibility of generative AI for advancing clinically-actionable tools that can improve cardiovascular disease phenotyping and diagnosis in real-world settings.

---

The electronic health record (EHR) is a powerful resource for patient-level research studying real-world implications of clinical data. Medical system-wide EHRs often capture data from patients across the socioeconomic spectrum including urban and rural areas, thus minimizing referral or selection bias. While rich with diverse information, the EHR also includes unstructured data and data missingness. Single level large language models (LLMs) have been proposed to improve extraction of EHR data; however, this approach is insufficient for phenotyping complex diseases and does not address the problem of data missingness.

Pulmonary hypertension (PH) is a highly morbid disease diagnosed by mean pulmonary artery pressure (mPAP) >20 mmHg during right heart catheterization (RHC)^1^. Since PH is a heterogenous pathophenotype, population studies are necessary to optimize mPAP clinical risk models.^2,3^ Large RHC cohorts (>10,000 patients) provide adequate statistical power for achieving this goal, and have already been used to clarify the PH hemodynamic clinical risk continuum. However, these datasets are subject to expert center referral bias. Furthermore, mPAP missingness is common in RHC cohort studies, again suggesting that mPAP risk models in use clinically may be subject to substantial bias. Although formulae have been proposed to impute mPAP based on available systolic PAP (sPAP) or diastolic PAP (dPAP), these approaches are derived from small ultra-selected patient subgroups^4-6^ limiting their generalizability.

We analyzed RHC reports (January 1, 2016 to December 31, 2024) from the University of Maryland Medical System (UMMS), a Maryland statewide clinical network serving > 2 million patients that includes 12 hospitals. Baseline demographics, including age, self-reported race and sex, height, weight, body mass index, date of RHC, and clinical comorbidities based on *International Classification of Disease-10 (ICD-10)* codes within 6 months of RHC^2,3^ were compiled.

We developed an automated LLM-driven Pattern Repository (LDPR) method using an 80/20-train-test split for extracting unstructured RHC data from the EHR at UMMS. The LDPR employs an agentic, multi-LLM architecture featuring three task-specific agents built on the quantized LLaMA 3.1 (70B) model. In the first step, the initial LLM agent determined whether a PAP value has been reported and extracts relevant segments of the whole RHC procedure note. A second LLM agent then validates these extracted segments and constructs RegEx-style patterns to formulize them. Finally, a third LLM agent applies and verifies the pattern against the original RHC procedure note to ensure alignment and accuracy of the defined pattern. These steps form an iterative algorithm, continuously expanding the LDPR. The finalized LDPR model was then cross-validated manually, independently by two experienced PH clinicians (K.Z., M.E.M.) on 500 reports randomly chosen overtime and across hospitals, allowing a 95% confidence interval and margin error of 5%.

In a subsequent analysis, we used the extracted sPAP and dPAP to develop a ML-derived mPAP calculation formula. We applied an 80/20 train-test split to the now structured dataset and developed both linear and polynomial regression models using sPAP alone or sPAP/dPAP combinations to predict mPAP. Next, model performance was evaluated using R2 and mean square error (MSE) and compared results against classic formulas using two-tailed t-tests. Model development and statistical testing were conducted in Python using the scikit-learn and scipy.stats packages. The study was approved by the University of Maryland Institutional Review Board.

The LDPR was developed and validated on 17,292 RHC reports from N=11,029 unique UMMS patients (mean [standard deviation] age, 66 [13.5] years, 43% female, 65% White, 30% Black, body mass index, 30 [7.7] kg/m2) enriched with typical cardiopulmonary diseases (26% congestive heart failure). The average mPAP in the cohort was 28 mmHg with a range of 4-94 mmHg. The precision for accurate mPAP, sPAP, and dPAP extraction (against manual adjudication) using the LDPR was 99.6%, 99.4%, and 99.4%, respectively. The precision for separating baseline PAP values from exercise, vasoreactivity testing, or confrontational fluid challenge (maneuvers performed during RHC in some patients) was 100%, 99.6% and 100%, respectively (Figure).

Detection failure for a missing (but available) PAP value was 0.4%. A missing mPAP, with available sPAP and dPAP, was noted in N=548 cases and N= 507 unique patients (3.2% and 4.6%, respectively). When applying ML to the dataset using linear and polynomial formulas, a simple, linear ML-derived equation (mPAP = 1.51 + 0.43*sPAP + 0.45*dPAP) achieved the highest R2 of 0.94 and lowest MSE of 8.3 mmHg. This formula outperformed mPAP prediction compared to previous classic linear equations^4-6^ (Figure). More complex polynomial regression derived by ML were not superior to the linear formula (p=0.47). When applying the ML-derived formula, 382 out of the 507 patients (75.3%) had a mPAP > 20 mmHg diagnostic for PH.

In our study, we demonstrated that the agentic, multi-LLM architecture extracted PAP data from a large, diverse, multi-center EHR with high accuracy in a training and validation cohort, demonstrating the effectiveness and scalability of this approach. Further, we show that a simple, linear ML-derived formula significantly outperformed classic equations to predict mPAP, highlighting the clinically actionable role of artificial intelligence in cardiopulmonary medicine.

RHC is the gold standard to diagnose, classify and risk stratify PH^1^ and is performed in PH expert and non-expert centers. Most published reports originate from PH expert centers, which can result in substantial reporting bias and underrepresentation of minority groups in the literature. Here, the EHR emerges as a critical tool for understanding real-world clinical management including accurate diagnosis^7^ by analyzing data from all patients that encounter a decentralized health care system. Despite this strong advantage of the EHR, sizable heterogeneity and missingness of embedded data challenge completeness and accuracy of EHR-based observations. The LLM overcomes the former by transforming data into a structured and uniform format, which allows for direct statistical usage. The development of an agentic LLM, as done in this work, is superior over a conventional LLM, as an agentic approach is able to work autonomously, therefore taking initiative without human programmer input, is goal-directed, maintains an internal memory and is adaptive, allowing change action mid-task if needed, all of which are not available by conventional LLMs.

Data missingness is a major problem for contextualizing research results to real world practice, since in the case of PH, diagnosis hinges directly on an available mPAP. In fact, in our data set, the imputation method reclassified 382 patients from non-diagnostic to PH, thereby influencing longitudinal prognosis and potentially qualifying them for treatment or PH-specific clinical research opportunities. In other published datasets, the missingness of mPAP from RHC reports was even higher with up to 7-10%^2,3^.

There are several classic formulae used to calculate a missing PAP. The first analysis by Chemla et al. (2004) was based on n=31 study subjects (22 with PH and 9 controls), including test and validation samples of 16 and 15 subjects, respectively^4^. Later, Chemla et al. (2009) used a total of n=166 individuals (n=94 with PH) to test additional linear formulae^5^, limiting their generalizability to large diverse cohorts despite the use of high-fidelity PAP measurements. Correspondingly, the MSE in our cohort ranged widely from 13 to 86 mmHg. To address this concern, we applied ML and derived a simple, linear equation that reduced the MSE to 8 mmHg, providing an accurate, scalable method to impute missing mPAP values, and enhance PH diagnosis and risk stratification in large EHR based RHC populations.

Our analysis is constrained by a retrospective design and potential variability in EHR documentation; however, these limitations seemed to be offset by our novel agentic, multi-LLM extraction approach and the large, diverse cohort that included > 17,000 RHC reports.

In conclusion, an agentic multi-LLM pipeline reliably structures RHC data and extracts PAP with high accuracy to provide an effective and scalable approach for PH research. A simple, linear ML-derived formula outperformed classic equations to predict mPAP, which may serve as a time- and cost-effective, widely deployable and highly accurate solution to the missing variable dilemma that confounds research studies and complicates clinical practice.

**Figure:**
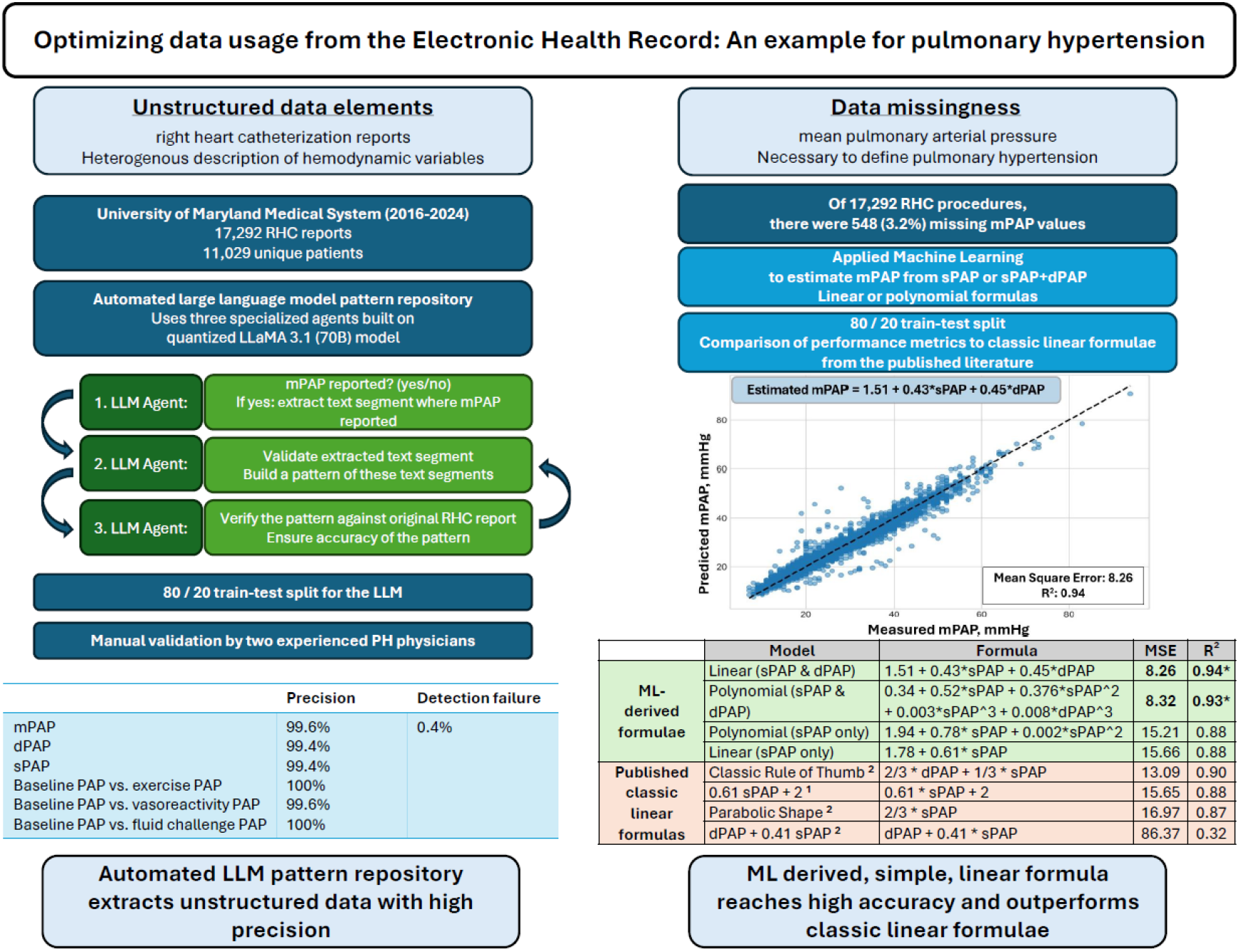
Optimizing data usage from the Electronic Health Record. Unstructured data elements can be extracted from the EHR with high accuracy by employing an automated large language model pattern repository. Data missingness in the EHR may be solved by Machine Learning derived imputation of missing variables. A ML-derived formula for predicting mean pulmonary arterial pressure (mPAP) yielded high accuracy and model performance metrics significantly improved previous classic linear formulae. Ref 1: Chemla et al. Chest 2004 Oct;126(4):1313-7 Ref 2: Chemla et al. Chest 2009 Mar;135(3):760-76 MSE: mean square error; sPAP: systolic PAP; dPAP: diastolic PAP.

## Data Availability

All data produced in the present study are available upon reasonable request to the authors and after ethical approval.

## Acknowledgements

We thank Ms. Chaitrali Kher and Mrs. Pratima Kshetry for excellent work with the electronic health record.

## Notes

**Funding:** The University of Maryland-Institute for Health Computing is supported by funding from Montgomery County, Maryland and The University of Maryland Strategic Partnership: MPowering the State, a formal collaboration between the University of Maryland, College Park, the University of Maryland, Baltimore, and the University of Maryland Medical System.

**Conflict of interest:** S.M.S, M.E.M., S.C., G.R. and C.M.E. report no conflicts. K.Z. reports research grants from United Therapeutics and Cardiovascular Medical Research and Education Fund (CMREF) and advisory board fees from AstraZeneca. B.A.M. has received grants from Deerfield Company; has a patent PCT/US2019/ 059890 pending to None, a patent #9,605,047 issued to None, a patent PCT/US2020/066886 pending to None, and a patent BWH 2023–152-29618-0438P02 pending to None; has received 5R01HL139613-03 grant, National Institutes of Health grant R01HL163960, National Institutes of Health grant R01HL153502, and National Institutes of Health grant R01HL155096-01.

### Competing Interest Statement

S.M.S, M.E.M., S.C., G.R. and C.M.E. report no conflicts. K.Z. reports research grants from United Therapeutics and Cardiovascular Medical Research and Education Fund (CMREF) and advisory board fees from AstraZeneca. B.A.M. has received grants from Deerfield Company; has a patent PCT/US2019/ 059890 pending to None, a patent #9,605,047 issued to None, a patent PCT/US2020/066886 pending to None, and a patent BWH 2023/152/29618/0438P02 pending to None; has received 5R01HL139613/03 grant, National Institutes of Health grant R01HL163960, National Institutes of Health grant R01HL153502, and National Institutes of Health grant R01HL155096/01.

### Funding Statement

The University of Maryland-Institute for Health Computing is supported by funding from Montgomery County, Maryland and The University of Maryland Strategic Partnership: MPowering the State, a formal collaboration between the University of Maryland, College Park, the University of Maryland, Baltimore, and the University of Maryland Medical System.

### Author Declarations

The study was approved by the University of Maryland Institutional Review Board and requirement for informed consent was waived.

